# Characterization of Circular RNA Transcriptomes in Psoriasis and Atopic Dermatitis Reveals Disease-specific Expression Profiles

**DOI:** 10.1101/2020.05.26.20090019

**Authors:** Liviu Ionut Moldovan, Lam Alex Tsoi, Stephen Weidinger, Johann Gudjonsson, Jørgen Kjems, Lasse Sommer Kristensen

## Abstract

**Background:** Atopic dermatitis (AD) and psoriasis, two chronic inflammatory skin diseases, affect a large number of individuals worldwide, and are associated with various comorbidities. Circular RNA (circRNA) constitute a major class of non-coding RNAs that have been implicated in many human diseases, although their potential involvement in inflammatory skin diseases remains elusive.

**Objectives:** To compare and contrast the circRNA expression landscapes in paired lesional and non-lesional skin from psoriasis and AD patients relative to skin from unaffected individuals. Moreover, to correlate circRNA expression to disease severity.

**Methods:** We analyzed high-depth RNA-seq data from paired lesional and non-lesional skin samples from 27 AD patients, 28 psoriasis patients, and 38 healthy controls. CircRNAs and their cognate linear transcripts were quantified using the circRNA detection algorithm, CIRI2.

**Results:** We identified 39,286 unique circRNAs in total and found that psoriasis and AD lesional skin could be distinguished from non-lesional and healthy skin based on circRNA expression landscapes. In general, circRNAs were less abundant in lesional relative to non-lesional and healthy skin. Differential expression analyses revealed many significantly downregulated circRNAs, mainly in psoriasis lesional skin, and a strong correlation between psoriasis and AD-related circRNA expression changes was observed. A subset of circRNAs, including ciRS-7, was specifically dysregulated in psoriasis and show promise as biomarkers for discriminating AD from psoriasis.

**Conclusion:** Psoriasis and circRNA transcriptomes share expression features, including a global downregulation, but only psoriasis is characterized by several circRNAs being dysregulated independently of their cognate linear transcripts.

## Introduction

Atopic dermatitis (AD) and psoriasis are the two most common chronic inflammatory skin conditions associated with a reduced quality of life and various comorbidities ^1, 2^. Both diseases derive from complex interactions of genetic and environmental factors and share features such as abnormal epidermal architecture, impaired keratinocyte differentiation and pronounced infiltration of inflammatory cells to the dermal layer. However, disease-specific molecular and cellular features distinguish the two diseases ^3, 4^. Several high-throughput RNA-sequencing (RNA-seq)-based studies have identified many dysregulated coding and non-coding RNAs in both psoriasis and AD ^5, 6, 7, 8^. Despite their similarities, they have distinct clinical presentations, genetic components, as well as molecular and cellular features ^8, 9, 10^.

Circular RNAs (circRNAs) form a large class of non-coding RNAs that are characterized by a covalent bond linking a downstream splice-donor site to an upstream splice-acceptor site ^11^. CircRNAs were previously viewed as products of aberrant RNA splicing events ^12^; however, recent research has revealed that circRNAs are highly abundant and have tissue and cell-type-specific expression patterns ^13^. Furthermore, many circRNAs are conserved across species ^14^ and some play important roles in a wide range of physiological and disease processes, including neurogenesis ^15^, cardiovascular diseases ^16^, diabetes mellitus ^17^, chronic inflammatory diseases ^7, 18^ and cancer ^19, 20^. Moreover, a recent study has reported that circRNAs are involved in the regulation of the innate immune system by binding and inhibiting the double-stranded RNA (dsRNA)-activated protein kinase (PKR). Upon poly(I:C) stimulation or viral infection, circular RNAs were globally degraded by RNase L, leading to the activation of PKR upon release ^18^. The same study provided a link between the global downregulation of circRNAs in patient-derived peripheral blood mononuclear cells and the inflammatory component of Systemic Lupus Erythematosus ^18^. Whether circRNAs may have similar functions in other chronic inflammatory diseases, including psoriasis and AD, has not been investigated. We recently reported a global downregulation of circRNAs in psoriasis lesional skin compared to non-lesional skin and healthy skin in a relatively small patient cohort ^7^, but circRNA transcriptomes in AD have not previously been profiled.

To gain a deeper understanding of the molecular changes associated with inflammatory skin diseases, we analyzed the circRNA transcriptomes within lesional and non-lesional skin biopsies from well-defined, large cohorts of AD and psoriasis patients, as well as skin biopsies from matched healthy control individuals. In total, we analyzed high-depth RNA-seq data from 135 samples, which were collected within an ongoing investigator-initiated clinical study ^8^. CircRNA expression changes were correlated with expression changes of their cognate linear host genes as circRNAs that are dysregulated independently of their respective host genes are more likely to be functionally relevant. Finally, we correlated circRNA expression changes with clinical scores (disease severity). Together, our analyses reveal differences and similarities of the circRNA expression landscapes and highlight key circular RNAs that may be directly involved in psoriasis and AD pathogenesis.

## Materials and Methods

### Study population

In this study, we analyzed high-depth RNA-seq data from 135 samples produced in a previous study ^8^. Informed written consent was obtained from human subjects under a protocol approved by the local ethics board at the University Hospital Schleswig-Holstein, Kiel, Germany (reference: A100/12). Adult patients with a history of AD or psoriasis for at least 3 years, as well as adult volunteers without a personal or familial history of atopic and chronic-inflammatory diseases, were invited to participate. Inclusion criteria for patients were a dermatologist-confirmed diagnosis of active chronic plaque-type psoriasis or AD. Exclusion criteria were the presence of any other chronic skin disease, systemic treatment with immune-efficient medication ever, and topical treatment within one week prior to material sampling. AD or psoriasis was diagnosed based on a skin examination by experienced dermatologists according to standard criteria (for AD, the American Academy of Dermatology Consensus Criteria were used) ^4, 21^. Table 1 offers a cohort description. For a detailed description of each patient see ^8^.

**Table 1.** Cohort description. Continuous traits as mean and standard deviation for atopic dermatitis patients (AD), psoriasis patients (Pso) and healthy control individuals (Healthy). Filaggrin (FLG) status (R501X, 2282del4, R2447X, S3247X) determined in AD and healthy control individuals only.

|  |  | AD | Pso | Healthy |
| --- | --- | --- | --- | --- |
| Number of individuals (male/female) |  | 27 (17/10) | 28 (14/14) | 38 (16/22) |
| Age |  | 34.07+/-10.96 | 41.89+/-15.74 | 32.63+/-11.64 |
| Objective Scord / PASI |  | 31.11+/-10.96 | 9.54+/-3.49 | - |
| Average Scord / PASI | Females | 25.97 +/- 9.73 | 4.32+/-2.53 |  |
|  | Males | 34.13+/-10.74 | 7.32+/-5.28 |  |
| BMI |  | 25.07+/-5.1 | 28.65+/-5.69 | 24.16+/-4.22 |
| FLG mutation carriers N (%) |  | 7 (25.93) | NA | 1 (2.63) |

### Sample preparation

Sequencing libraries were prepared from total RNA extracted from 135 skin biopsies (38 control; 27 psoriasis non-lesional (NL); 28 psoriasis lesional (LL); 21 AD NL; 21 AD LL). The preparation methods and quality control procedures have been described ^8^. Briefly, total RNA was isolated from PAXgene fixed tissue samples using the AllPrep DNA/RNA Mini Kit (Qiagen, Hilden, Germany) following the manufacturer’s specifications. Only samples with a concentration of >50 ng/μl, an optical density 260/280 ≥1.8, and an RNA integrity number >7 were included in subsequent library preparation and sequencing.

### RNA library preparation and sequencing

RNA samples were prepared for sequencing using the Illumina Truseq® Stranded total RNA Protocol in combination with the RiboZero rRNA removal Kit and sequenced on the HiSeq2500 in pools of 10 samples with 2×125bp according to the manufacturer’s protocol (Illumina, San Diego, CA) ^8^. On average 91 million paired-end reads were obtained per sample. The samples were randomly distributed across the plates and pools before sequencing to minimize batch effects.

### circRNA analyses in RNA-seq data

Raw data were downloaded from the ENA database. Sequencing reads were quality filtered, trimmed and adaptor sequences were removed using trim_galore (v0.4.1). CircRNAs were quantified in all samples individually using CIRI2 ^22^. Raw back splice-junction (BSJ)-spanning reads were analyzed using DEseq2 for normalization (median of ratios) and differential expression analysis (using a false discovery rate of <0.05 and |log2 (fold change) | >1) to identify differentially expressed (DE) high-abundance circRNAs (having an average of at least five back splice-junction (BSJ)-spanning reads among all samples) among the different sample groups. Four samples were removed from the analysis due to a relatively low amount of overall data.

### Statistical analyses

All statistical tests were performed using Prism 7 (GraphPad, La Jolla, CA, USA). Correlation analyses were done using Spearman correlation analysis for non-normal data. The hybrid Wilson/Brown method was used to determine the confidence intervals of the proportions when constructing receiver operating curves. Comparisons of the average expression levels of the high-abundance circRNAs between the lesional-, non-lesional and healthy skin were done using the Wilcoxon test for matched data and the Mann-Whitney test for unmatched data respectively, as the data were not normally distributed according to the D’Agostino & Pearson normality test. All P-values were two-tailed and considered significant if < 0.05.

## Results

### CircRNA transcriptomic architecture of psoriasis- and AD lesional and non-lesional skin

By analyzing high-depth RNA-seq data, we performed an unbiased and genome-wide profiling of circRNAs in patients suffering from psoriasis and AD and compared the findings with the profiles of skin from healthy controls. We analyzed paired lesional and non-lesional skin samples from 21 AD patients and 28 psoriasis patients. Additionally, 38 healthy control subjects were included in the study. In the entire dataset, we detected 39,286 circRNAs supported by at least two BSJ spanning-reads in a single sample using the CIRI2 algorithm ^22^ (Table S1). Interestingly, when performing principal component analysis (PCA) using all circRNAs identified, we found that the first two principal components could clearly separate the healthy control skin from psoriasis and AD lesional skin, with psoriasis lesional skin samples showing the most distinct profile. Moreover, the non-lesional skin samples from psoriasis and AD patients showed profiles extending towards the lesional samples relative to the healthy skin samples, although many non-lesional skin samples overlapped with healthy skin samples (Figure 1a). Interestingly, we also observed that the non-lesional skin samples that were most distinct from the healthy skin samples were from male patients. This applied to both psoriasis and AD (Figure 1a).

**Figure 1.**
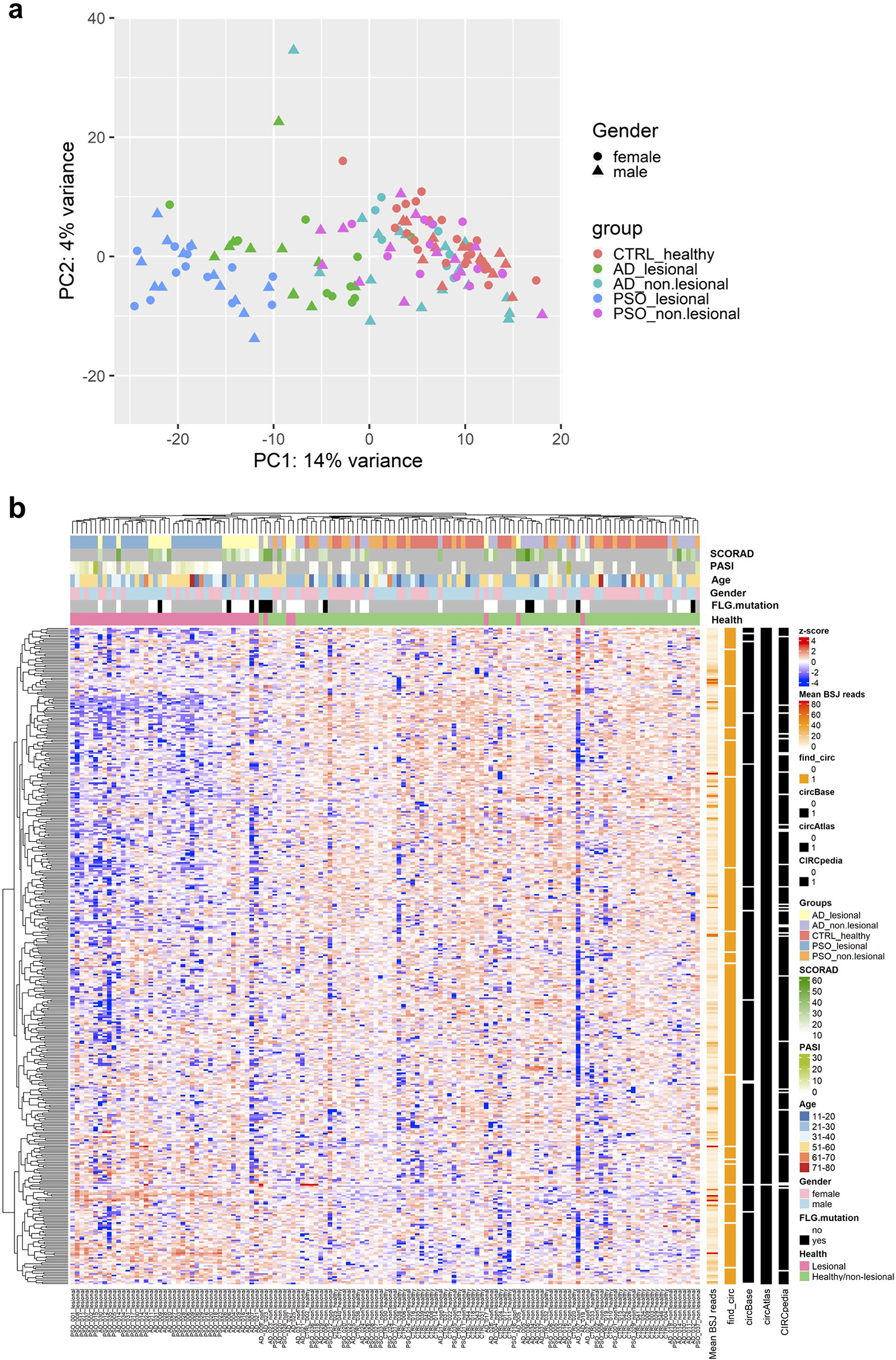
Comparative analysis of the circRNA and protein-coding transcriptome in psoriasis and atopic dermatitis. (a) Principal component analysis of circRNA expression in samples obtained from healthy and paired lesional and non-lesional psoriasis and AD skin, based on RNA-seq data. (b) Heatmap and hierarchical clustering of 414 high-abundance circRNAs (supported by an average of at least five BSJ-spanning reads among all samples).

Next, we focused on the most abundant circRNAs, rationalizing that these are more likely to be of functional relevance. Thus, we defined a high-abundance set consisting of 414 circRNAs that were supported by an average of at least five BSJ-spanning reads among all samples (Table S2). To visualize circRNA expression patterns in individual samples, we displayed the data from the high-abundance circRNAs as a heatmap with associated hierarchical cluster analyses on both axes (Figure 1b). Again, we observed that lesional skin samples generally clustered separately from non-lesional and healthy skin samples, mainly driven by downregulation of a large number of circRNAs in the lesional samples. On the other hand, the clinical parameters considered (age, gender, *FLG* mutation and clinical scores (SCORAD and PASI)) did not seem to have an impact on the clustering.

### Identification of differentially expressed circRNAs between the different sample groups

Differential expression analyses (using a false discovery rate of <0.05 and |log2 (fold change) | >1) were conducted to identify differentially expressed (DE) high-abundance circRNAs among the different sample groups (Figure 2, Figure S1 and Table S3). The majority of the DE circRNAs were downregulated in psoriasis lesional skin relative to healthy skin (Figure 2a, b) and, overall, circRNAs were significantly less abundant in the lesional skin relative to healthy and non-lesional skin (Figure S2a). In total, 60 circRNAs were significantly downregulated by more than 2-fold, most significantly ciRS-7, circEXOC6B, circSLC8A1, and circRHOBTB3, whereas 11 circRNAs were upregulated by more than 2-fold, most significantly circTNFRSF21, circDOCK1, circARAP2, circZRANB1 and circDDX21 (Figure 2b). When comparing AD lesional skin to healthy control skin, we observed fewer DE circRNAs, but again most were downregulated in the lesional skin (Figure 2a, c) and the overall circRNA expression was significantly reduced relative to non-lesional and healthy skin (Figure S2b). In total, 10 circRNAs were significantly downregulated by more than 2-fold, most significantly circRHOBTB3, circDEGS1, circDEF6, circSWT1, and circCCDC7, whereas two circRNAs were significantly upregulated by more than 2-fold (circTNFRSF21 and circDDX21). By comparing AD and psoriasis lesional skin, we found that the majority of the DE circRNAs to be more abundant in the AD samples (Figure 2a, d). In particular, we observed that ciRS-7 was much more abundant in AD lesional skin relative to psoriasis lesional skin. Finally, we compared the non-lesional skin of the two conditions with the healthy skin, but found no DE circRNAs (Figure S1). However, we found it interesting that ciRS-7 and circTNFRSF21 were the most downregulated and upregulated circRNAs, respectively, in non-lesional psoriasis skin relative to healthy skin, consistent with what we observed in the lesional skin.

**Figure 2.**
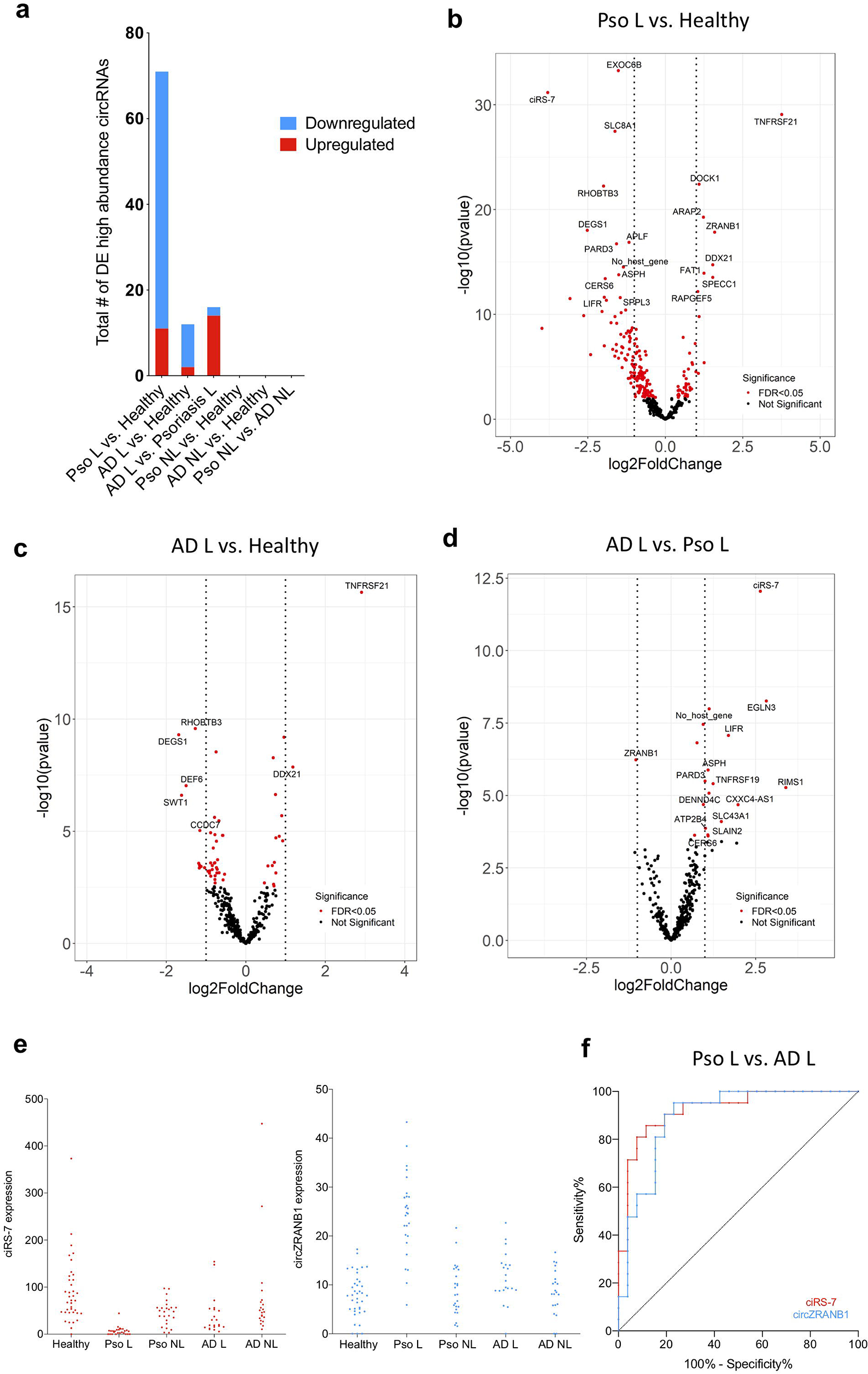
Differential expression analysis of the high abundance circRNAs among the different sample groups. (a) Number of differentially expressed high abundance circRNAs in lesional and non-lesional psoriasis and AD skin, relative to healthy controls. Volcano plots showing the differential expression analyses of all high abundance circRNAs (log2 fold change versus -log10(P-value) comparing psoriasis lesional skin relative to healthy skin (b), AD lesional skin relative to healthy skin (c) and lesional skin in psoriasis and AD (d). Vertical lines denote the |log2 (fold change)|>1. Red colour represents DE high abundance circRNAs with a false discovery rate of <0.05. (e) Scatter plots illustrating the expression distributions of ciRS-7 and circZRANB1 across the different skin conditions. (f) Area under receiver operating characteristic to evaluate the performance of classifying Pso L and AD L skin using the ciRS-7 and circZRANB1 expression profiles. The hybrid Wilson/Brown method was used to determine the confidence intervals of the proportions.

### ciRS-7 and circZRANB1 are promising diagnostic biomarkers

Despite distinct clinical features of AD and psoriasis, they have a significant overlap in their molecular architecture and their differential diagnosis can sometimes be problematic. Therefore, as we observed that some circRNAs (circEGLN3, circLIFR, circRIMS1, circASPH, circPARD3, circTNFRSF19, circCXXC4-AS1, circDENND4C, circSLC43A1, circATP2B4, circSLAIN2, circCERS6) were differentially expressed between AD lesional and psoriasis lesional skin samples, we investigated their potential as diagnostic biomarkers by constructing receiver operating curves for each; area under the curve was 0.82 for circEGLN3, 0.84 for circLIFR, 0.72 for circRIMS1, 0.70 for circASPH, 0.73 for circPARD3, 0.75 for circTNFRSF19, 0.78 for circCXXC4-AS1, 0.79 for circDENND4C, 0.71 for circSLC43A1, 0.78 for circATP2B4, 0.86 for circSLAIN2 and 0.75 for circCERS6. In particular, ciRS-7, which was much less abundant in psoriasis, and circZRANB1, the only circRNA significantly higher in psoriasis lesional skin relative to AD lesional skin, exhibited the biggest potential as diagnostic biomarkers (the area under the curve was 0.92 for ciRS-7 and 0.89 for circZRANB1) (Figure 2e, f).

### A strong correlation between circRNA expression changes observed in psoriasis and AD

We found that nine out of ten downregulated circRNAs in AD lesional skin relative to healthy skin (ciRS-7, circRHOBTB3, circDEGS1, circDEF6, circCCDC7, circCCDC7, circPOF1B, circSUMF1, circSWT1) were also all downregulated in psoriasis lesional skin. Moreover, the only two upregulated circRNAs in AD lesional skin (circTNFRSF21 and circDDX21) were also upregulated in psoriasis lesional skin (Figure 3a). In line with these observations, we found a strong correlation between circRNA expression changes in psoriasis lesional skin and in AD lesional skin (relative to the healthy controls) (r = 0.78 and P < 1 × 10^-4^) (Figure 3b). Although there were no significant differences between psoriasis non-lesional skin and healthy skin, we observed a significant correlation between the circRNA expression changes in psoriasis non-lesional skin relative to healthy skin and the circRNA expression changes in psoriasis lesional skin relative to healthy skin (r = 0.40 and P < 1 × 10^-4^) (Figure 3c). This suggests that minor circRNA expression changes from healthy to non-lesional skin are further augmented in the lesional skin. Similarly, we observed a significant correlation between circRNA expression changes in AD non-lesional skin relative to healthy skin and the circRNA expression changes in AD lesional skin relative to healthy skin (r = 0.56 and P < 1 × 10^-4^) (Figure 3d).

**Figure 3.**
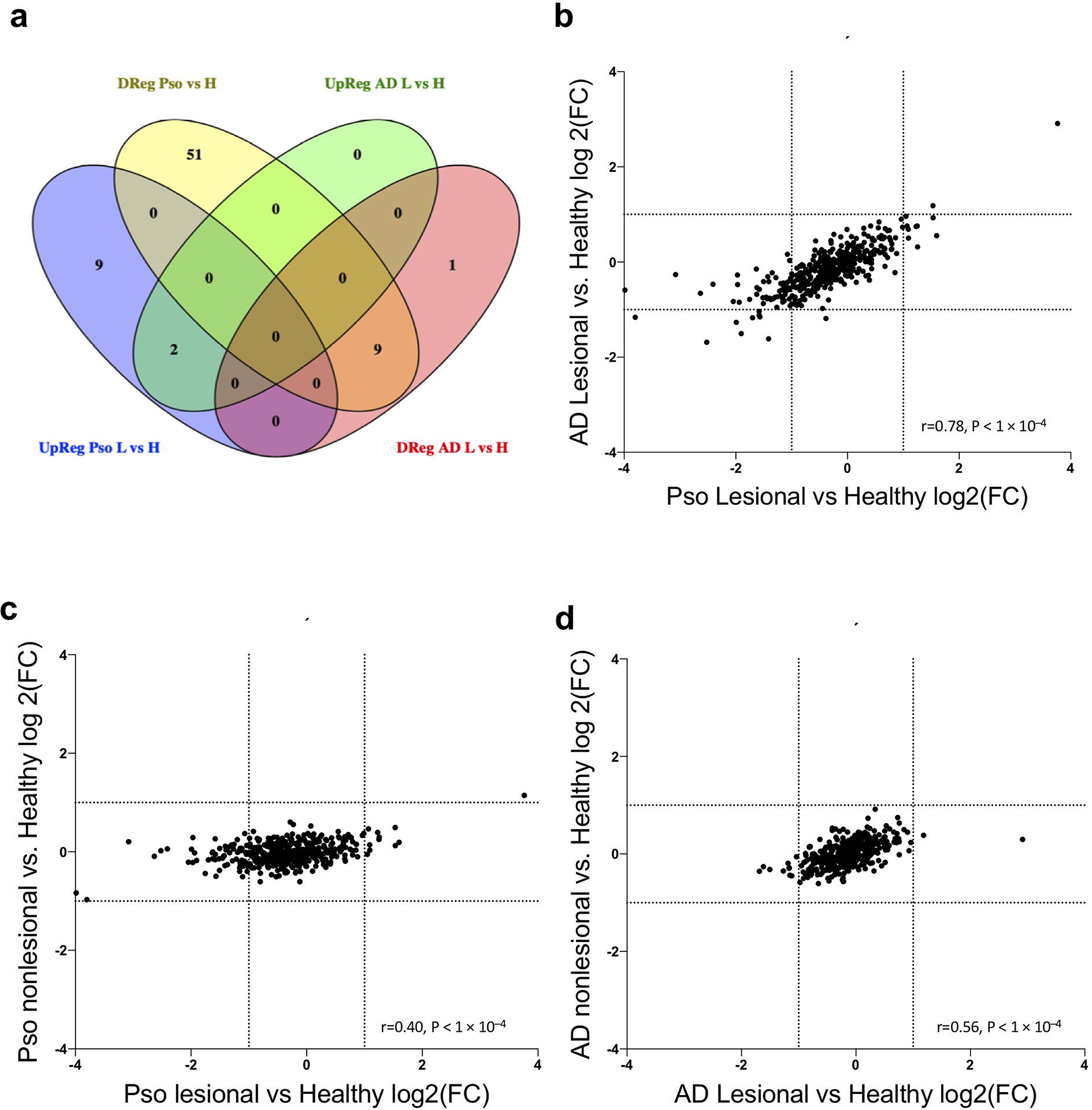
Correlation between circRNA expression changes observed in psoriasis and AD. (a) Venn diagram showing the overlap between the DE high abundance circRNAs in the lesional psoriasis and AD skin. Scatter plots illustrating the concordance between circRNA expression profiles in lesional skin for psoriasis and AD (b), and non-lesional and lesional psoriasis (c) and AD skin (d). Correlation analyses were done using Spearman correlation for non-normal data.

### Many DE circRNAs in psoriasis lesional skin are changed independently of their cognate linear transcripts

Next, we investigated whether the DE circRNAs identified in psoriasis and AD lesional skin were altered independently of their respective cognate linear transcripts. Therefore, we plotted circRNAs expression changes against the corresponding changes of their respective cognate linear transcripts (Figure 4). We observed a significant correlation between the circRNA and linear RNA expression changes, both in psoriasis and in AD, relative to healthy skin (r=0.44 and r=0.57, respectively, P < 1 × 10^-4^). However, in the case of psoriasis, there was also a substantial number of circRNAs (31 out of 66 downregulated circRNAs) that were expressed independently of their linear host genes. In particular, ciRS-7, circLIFR, circVCAN, circDEGS1, circCXXC4-AS1, circRIMS1 and circEGLN3 were all more than 4-fold downregulated in psoriasis lesional skin, while their cognate linear transcripts were relatively stable. Moreover, four circRNAs, two circDOCK1 isoforms, circZRANB1 and circGRAMD4 were more than 2-fold upregulated independently of the linear transcripts (Figure 4a). In AD lesional skin, however, no circRNAs were found to be dysregulated independently of their cognate linear transcripts (Figure 4b).

**Figure 4.**
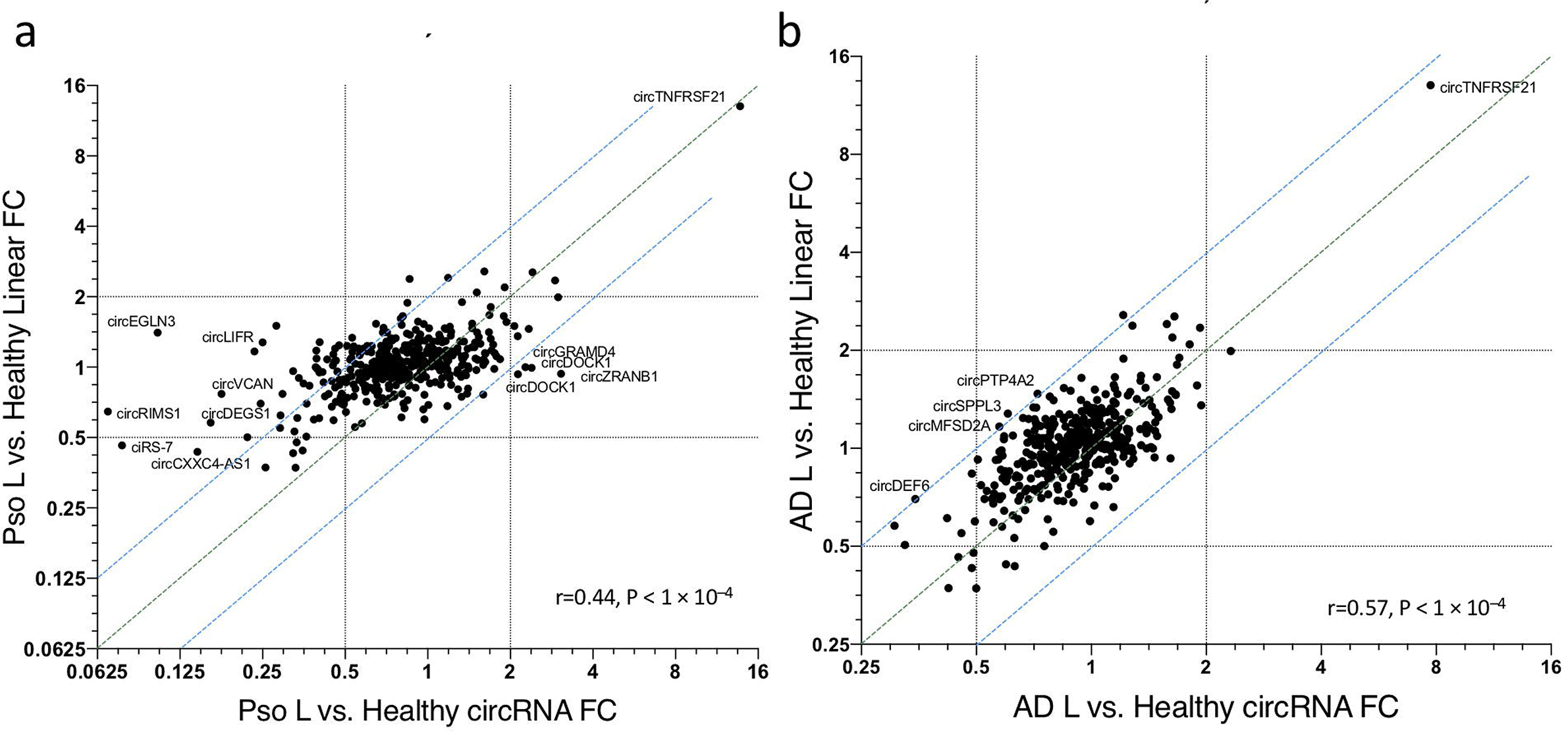
Correlation analysis between the expression changes of circRNAs and changes of their cognate linear host transcripts in lesional relative to healthy skin. The dotted blue lines are set at one fold change from the identity line represented in green and circRNAs found outside the blue lines are considered to be regulated independently of their linear host genes. Correlation analyses were done using Spearman correlation for non-normal data. (a) Psoriasis. (b) AD.

### circRNA analysis may have translational inference

Next, we analyzed whether changes in circRNA abundance correlate with the disease severity scores (PASI for psoriasis and SCORAD for AD). Firstly, we correlated the average expression levels of the high-abundance circRNAs in each lesional skin sample with the respective disease severity score (Figure S3a). Although not significant, we found a negative correlation both when considering PASI and SCORAD, which was expected, as circRNAs were globally downregulated in lesional skin relative to non-lesional skin. We then considered the average expression levels of circRNAs that were more than two-fold downregulated in the lesional skin (Figure S3b) and found the same negative correlations, slightly stronger in the case of AD, but still not significant. Finally, we investigated circRNAs that were downregulated both independently and dependently on their cognate linear host transcripts in the psoriasis lesional skin (where the effect was greater than in AD, as described above) (Figure S3c). While the correlations were minor and not significant, only the independent circRNAs correlated negatively with PASI score.

## Discussion

Psoriasis and AD have long been considered to exist on a ‘spectrum’ ^23^ based on common underlying immunologic processes, common treatment approaches ^24^ and the histological resemblance of chronic AD with psoriasis ^25^. Recently, transcriptomic studies have shed light on the common and unique molecular traits of both diseases ^8^ and while inflammation-related transcripts overlap to a certain degree in both lesional and non-lesional skin, features such as IL-13/IL-4 response in AD and IL-17 response in psoriasis clearly separate the two disease entities. Next-generation sequencing has facilitated the profiling and functional description of several classes of non-coding RNAs, such as long non-coding RNAs and miRNAs, in both psoriasis and AD ^6, 26, 27, 28, 29^, and was recently extended to circRNAs in psoriasis ^7, 30^. In contrast, circRNAs have not previously been investigated in AD.

Our deep-sequencing based study provides insights into the common and specific features of the circRNA transcriptomes of psoriasis and AD in both lesional and non-lesional skin. Using a *de novo* circRNA detection algorithm ^31^, CIRI2 ^22^, we identified 39,286 unique circRNAs in biopsies from lesional and non-lesional psoriasis and AD skin as well as healthy controls. In line with our previous findings ^7^ using RNA-sequencing and NanoString nCounter analysis ^32^, we observed a reduction of circRNA abundance in psoriasis lesional skin compared to non-lesional and normal skin. Interestingly, we observed the same phenomenon in AD, albeit less pronounced. Moreover, PCA and hierarchical cluster analyses revealed that the healthy control skin samples could be clearly separated from psoriasis and AD lesional skin samples based on circRNA expression data. Interestingly, we observed that the non-lesional skin samples from male patients were generally most distinct from the healthy skin samples, while also having a higher average severity score than female patients (Table 1).

A recent study showed that circRNAs are degraded by an endoribonuclease, RNase L, and that they are globally downregulated in autoimmune diseases such as Systemic Lupus Erythematosus ^18^. Therefore, we speculate that a similar mechanism could be responsible for circRNA regulation in inflammatory diseases such as psoriasis and AD. Alternatively, it is possible that the observed circRNA downregulation is due to the relatively slow turnover of circRNAs, which may prevent them from reaching steady-state levels ^33^ in the highly proliferative keratinocytes. Of note, circRNAs have also been found to be generally downregulated in the proliferative skin disease, cutaneous squamous cell carcinoma ^34^, however, the mechanism is not yet understood.

Many individual circRNAs were differentially expressed between lesional and healthy skin in both psoriasis and AD. Interestingly, when comparing to healthy skin, all significantly upregulated circRNAs in AD were also upregulated in psoriasis and nine out of ten downregulated circRNAs in AD were among the 60 significantly downregulated circRNAs in psoriasis. This points to similar mechanisms being active in the lesional skin of both disease processes. However, several circRNAs were differentially expressed between psoriasis lesional skin and AD lesional skin. In particular, ciRS-7 and circZRANB1 were the most significantly downregulated and upregulated circRNAs in psoriasis relative to AD, respectively, and exhibited strong potential as diagnostic biomarkers.

We observed a general correlation between changes in circRNA expression and their cognate linear host transcripts in both diseases; however, in psoriasis we found a number of circRNAs to be dysregulated independently of their respective host genes. Among these were ciRS-7 (also known as CDR1as), the most well-characterized member of the circRNA family ^14, 35^, which we previously found to be specifically downregulated in the epidermis (most pronounced in the basal stem cell layer) ^7^. While ciRS-7 is on average downregulated by 3.79-fold in psoriasis, it was not differentially expressed in AD. ciRS-7 was recently found to be silenced during melanoma progression and this loss of expression contributed to the metastatic spread of melanoma cells ^36^. Furthermore, through its association with miR-7, ciRS-7 was hypothesized to modulate *EGFR* and *KLF4* during breast cancer development, two genes that also play important roles in the skin ^37, 38^ Two other significantly downregulated circRNAs in psoriasis and AD, circRHOBTB3 and circSLC8A1, were also found to be downregulated in gastric and bladder cancer, respectively ^39, 40^. Moreover, their overexpression in cancer cells inhibited cell proliferation, suggesting that the observed downregulation of these circRNAs in both diseases may contribute to the pronounced keratinocyte proliferation. circTNFRSF21, known the be upregulated in cutaneous squamous cell carcinoma relative to healthy skin ^34^, was more than 4-fold upregulated in both psoriasis and AD lesional skin compared to healthy skin. The upregulation of circTNFRSF21 was, however, dependent on the expression of its host gene, *TNFRSF21*, which plays a role in inflammation and immune regulation ^41^. Finally, circZRANB1, which was significantly upregulated independent of its linear host transcript, was recently proposed as a promising therapeutic target in glaucoma-related retinal degeneration ^42^.

We also observed significant correlations between the circRNA expression differences in the lesional skin of the two diseases relative to healthy skin, as well as between the circRNA expression differences in non-lesional skin relative to lesional skin. This underscores the progression from healthy to uninvolved to lesional skin and describes an important degree of concordance between the circRNAome of the two conditions. Thus, we hypothesized that circRNA expression abundance would correlate negatively with disease severity. Both in psoriasis and AD, total circRNA abundance was negatively correlated with disease severity, although not statistically significant. Of note, in psoriasis only the independently downregulated circRNAs were negatively correlated with PASI, warranting future investigations into their potential roles in the disease pathogenesis. However, since most of the patients in our cohort had a mild and moderate disease phenotype, a more heterogeneous cohort in terms of PASI and SCORAD, involving patients with a more severe disease phenotype should be investigated to shed further light on possible correlations between circRNA expression and disease severity.

In summary, our study provides a comprehensive list of deregulated circRNAs in psoriasis and AD. These data are consistent with the notion that these diseases are to a certain extent overlapping at the molecular level, although each of them has distinct characteristics. The findings entail the need for future investigations to dissect the functional roles of circRNAs in these diseases, as well as how their expression patterns change after specific treatments and if circRNAs could be considered as diagnostic biomarkers or as an alternative for the evaluation of disease severity. Future research is warranted to chart their potential as therapeutic targets in psoriasis and AD, and to gain a better understanding of their functional roles in the molecular pathogenesis of these diseases.

## Data Availability

National Center for Biotechnology Information Gene Expression Omnibus accession: GSE121212.

## Acknowledgments

We thank Morten Venø for his outstanding bioinformatical assistance and Anne Færch Nielsen for a thorough and critical reading of the manuscript.

Funding sources: This project has received funding from the European Union’s Horizon 2020 research and innovation program under the Marie Skłodowska-Curie grant agreement No. 721890. The funding body provided financial support, but had no other role in the design of the study, data collection, analyses, and interpretation of data, decision to publish, or preparation of the manuscript.

---

**Figure S1.**
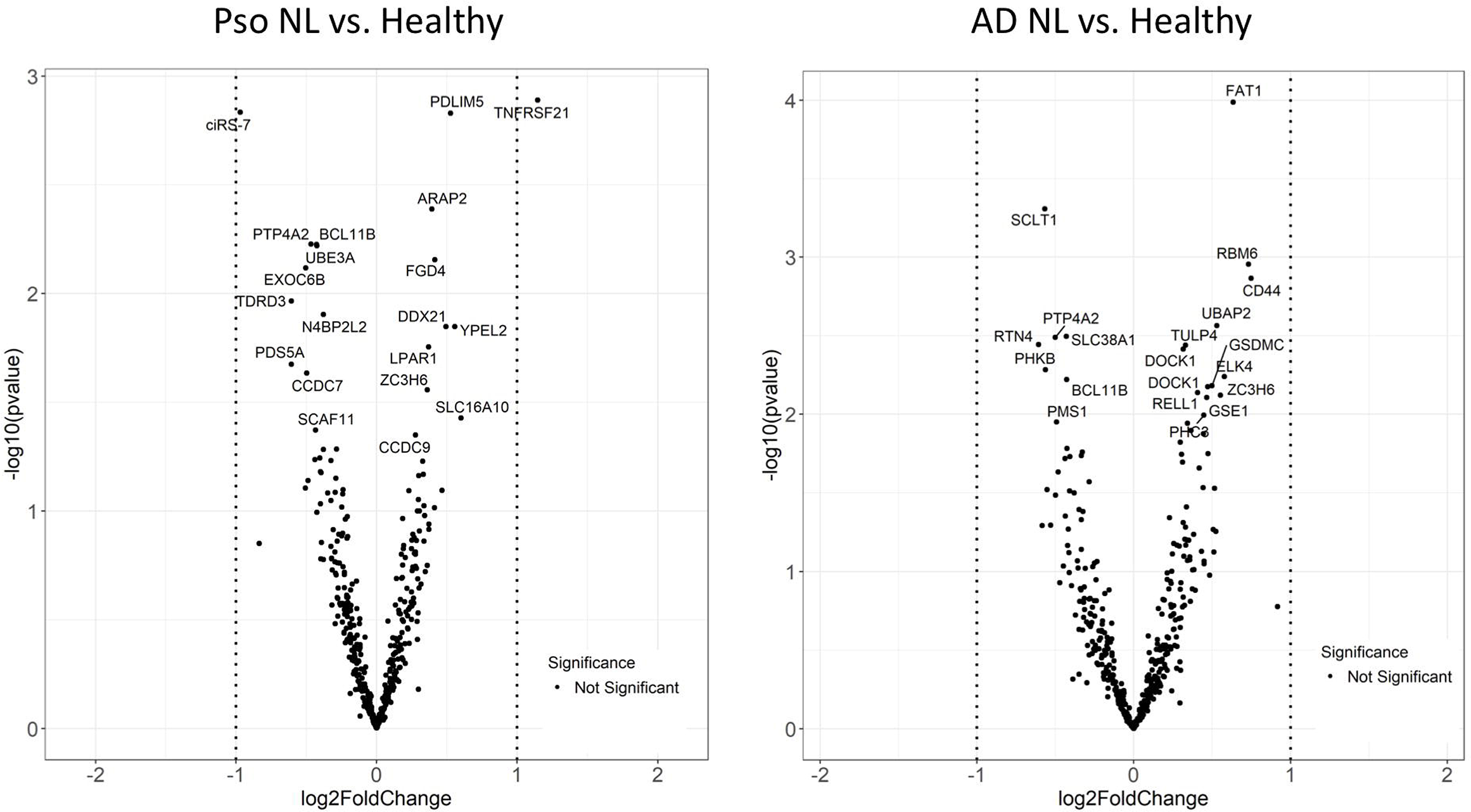
Differential expression analysis of the high-abundance circRNAs in non-lesional skin samples. Volcano plots showing the results of differential expression analyses of all high-abundance circRNAs (log2 fold change versus -log10(P-value) for all high-abundance circRNAs). Vertical lines denote a |log2 (fold change)|>1.

**Figure S2.**
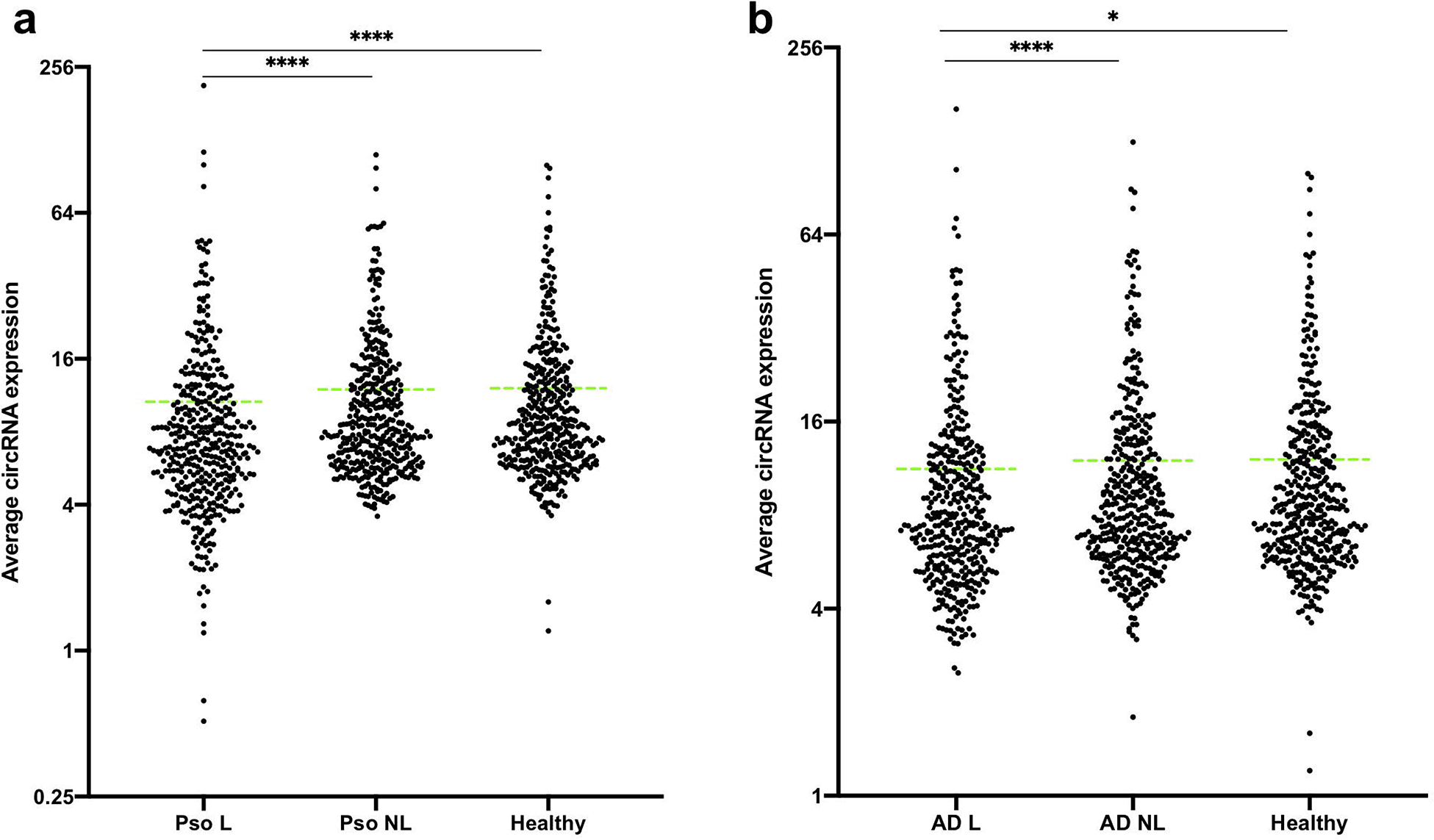
Column scatter plot showing the average high-abundance circRNAs expression detected in the normal and lesional and non-lesional skin biopsies combined. *P < 0.05, ****P <0.0001, Wilcoxon test for matched data (L vs. NL) and the Mann-Whitney test for unmatched data (L vs. Healthy).

**Figure S3.**
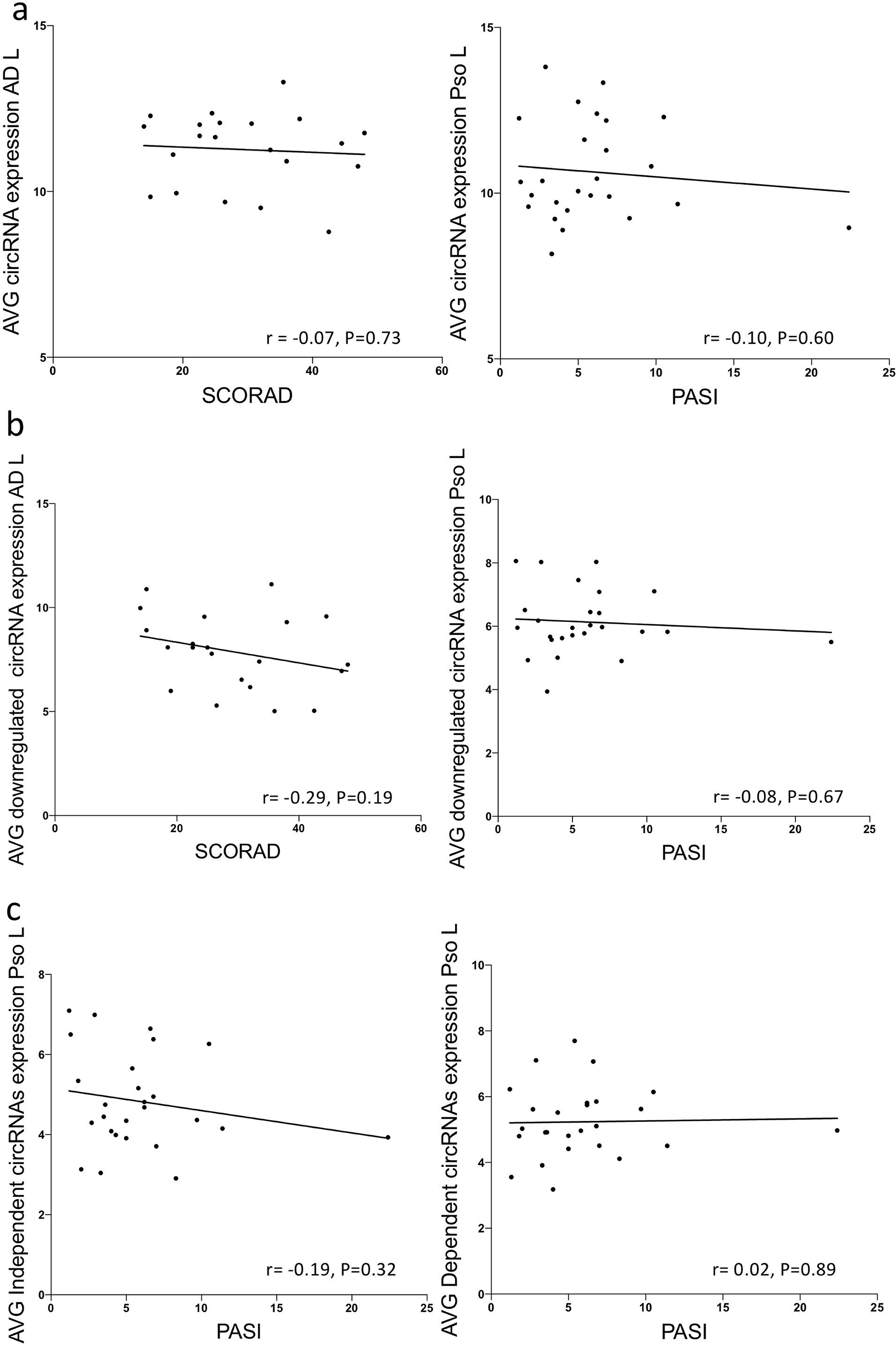
Associations between circRNA expression changes and the disease severity scores. Scatter plots illustrating the relation between the disease severity indexes PASI and SCORAD (x-axis) and the average expression levels of all 414 high-abundance circRNAs in each AD and psoriasis lesional skin sample (a), the average expression levels of all downregulated circRNAs in AD and psoriasis (b) and average expression levels of circRNAs that were more than two-fold downregulated in psoriasis lesional skin, both independently and dependently on their linear host genes (c) (y-axis). Correlation analyses were done using Spearman correlation for non-normal data.

